# A retrospective case - control study for assessing the risk factors for development of Diabetic Kidney Disease among people with Type 2 Diabetes in Tamil Nadu and Puducherry

**DOI:** 10.1101/2022.12.02.22283018

**Authors:** Sneha Saji, Srividya Suresh, Deepak MC, Sampath Kumar Krishnaswamy, Arthur Joseph Asirvatham, Manoj Kumar, Melvin George, Subramaniyan Kumaraswamy, Narayanan Krishnamoorthy, Gopinath Raman, Arun Kannan, Ratnesh Bhai Mehta

## Abstract

**Objective:** To understand the associated risk factors in the progression of Diabetic Kidney Disease among the Type 2 Diabetes individuals living in the state of Tamil Nadu and Puducherry.

**Research design and methods:** Clinical and socio-demographic data was collected, digitized, and analyzed for 482 participants diagnosed either with Diabetic Kidney Disease (DKD) or Type 2 Diabetes (T2D). The study was analyzed by using descriptive statistical analysis SAS version 9.4.

**Results:** Out of 482 participants, 422 fulfilled the eligibility criteria. Gender, age, T2D duration, and comorbidity are the major risk factors that are found to be associated with DKD in population understudy. We also found inclination towards usage of insulin among DKD participants rather than oral diabetic medications. Metformin (Biguanides) was the most used oral diabetic medication among the T2D participants followed by DPP-4 inhibitors and Sulphonylureas.

**Conclusion:** Together, these data describe the risk pattern among participants diagnosed with DKD at regional level that is integral in early and proper management of the disease.

## Introduction

Type 2 Diabetes Mellitus (T2D) is the most common form of diabetes, constituting 90% of the diabetic population worldwide (Wild et al. 2004) and the number has been steadily rising in low- and middle-income countries (Brown and for CDC 2009). Globally, prevalence of T2D has increased by ∼10-12% in the last two decades (Tewari et al. 2021). According to the Diabetes Atlas 2019, India has the largest number of diabetic patients in the world, estimated around 40.9 million in 2007 and expected is to increase by 69.9 million by 2025 (Unnikrishnan I et al. 2007). The impact of sedentary living, high-energy dietary intake and other unclear genetic and environmental factors are the main causes of rise in the prevalence and incidence of T2D (Kolb and Martin 2017). People diagnosed with T2D often have at least one additional comorbidity ranging from hypertension and obesity to kidney, liver disease and sleep apnea (Fruh 2017). The comorbidity burden increases with age and is higher in men compared to women (Iglay et al. 2016).

Diabetic Kidney Disease (DKD) alias diabetic nephropathy is a major long-term complication of T2D and is the leading cause of chronic kidney disease (CKD) and end stage kidney disease (ESRD) (Farah et al. 2021). In 2017 globally, there were 697.5 million CKD cases with one third living in China (132.3 million) and India (115.1 million) (Bikbov et al. 2020). Around 40% of the T2D diagnosed patients develop DKD which has a major effect not only on global health in respect to mortality, morbidity, but also puts burden on the family (Alicic et al. 2017). The factors contributing to increase in the risk of kidney disease in T2D patients include hypertension, uncontrolled blood sugar, obesity, presence of other comorbidities etc. (Hussain et al. 2021a).

Interestingly, DKD often stays undiagnosed until severe complication manifestation starts to show up (Hussain et al. 2021a). Many researchers have noticed that delayed diagnosis may have arisen from insufficient knowledge and understanding about kidney disease (Vassalotti et al. 2010). Considering variability across the globe in respect to prevalence and incidence of kidney disease among T2D patients and to implement region specific screening program in early course of disease, careful assessment of DKD epidemiology is needed.

There are few studies published on the prevalence of diabetic nephropathy in India (Kumar et al. 2022), (Tewari et al. 2021). Efforts have also been made to understand the prevalence, and determinants associated with CKD at regional level (Parameswaran et al. 2020; Bala et al. 2021; Eswarappa et al. 2022). Although attempts have been made in countries like India (Kumar et al. 2022), but still limited information is available around baseline characteristics of DKD not only at regional level but also at national level. Hussain and coworkers conducted the study to assess the awareness of kidney disease among T2D patients in India and found that 56% of them had poor information around kidney function (Hussain et al. 2019). A cross-sectional study conducted among the Tamil Nadu population reported that the participants had ample knowledge of the risk factors, signs, and symptoms of CKD, while insufficient knowledge on the kidney’s physiological function, and the diagnosis of CKD (Bala et al. 2021).

Current study has a refined set of participants from southern part of India primarily Tamil Nadu and Puducherry. It is a case-control observational study that tries to provide a baseline characteristic associated with epidemiological, clinical, and therapeutic management in DKD patients in the real world setting at regional level.

## Materials And Methods

### Study Design

The current study is a retrospective observational multicentric cohort study of people who have Type 2 Diabetes (T2D) and/or Diabetic Kidney Disease (DKD). Prior to participant recruitment, the study was approved by an independent institutional ethics committee (IEC) for each site and the study was registered with Clinical Trials Registry-India. The study and its associated procedures were carried out in accordance with the Helsinki Declaration guidelines.

A total of 500 participants were planned to be recruited across 3 cities, 9 centers around Tamil Nadu and border sharing union territory (Puducherry). Participants visiting clinics/hospitals for their outpatients’ visits such as routine checkup or undergoing dialysis who met the eligibility criteria were invited to participate in the study (Table 2). Based on their clinical history and biochemical test results, all the participants were divided into two groups: control (those with T2D who had a healthy kidney) and case (participants having T2D with decrease in kidney function).

**Table 1:**
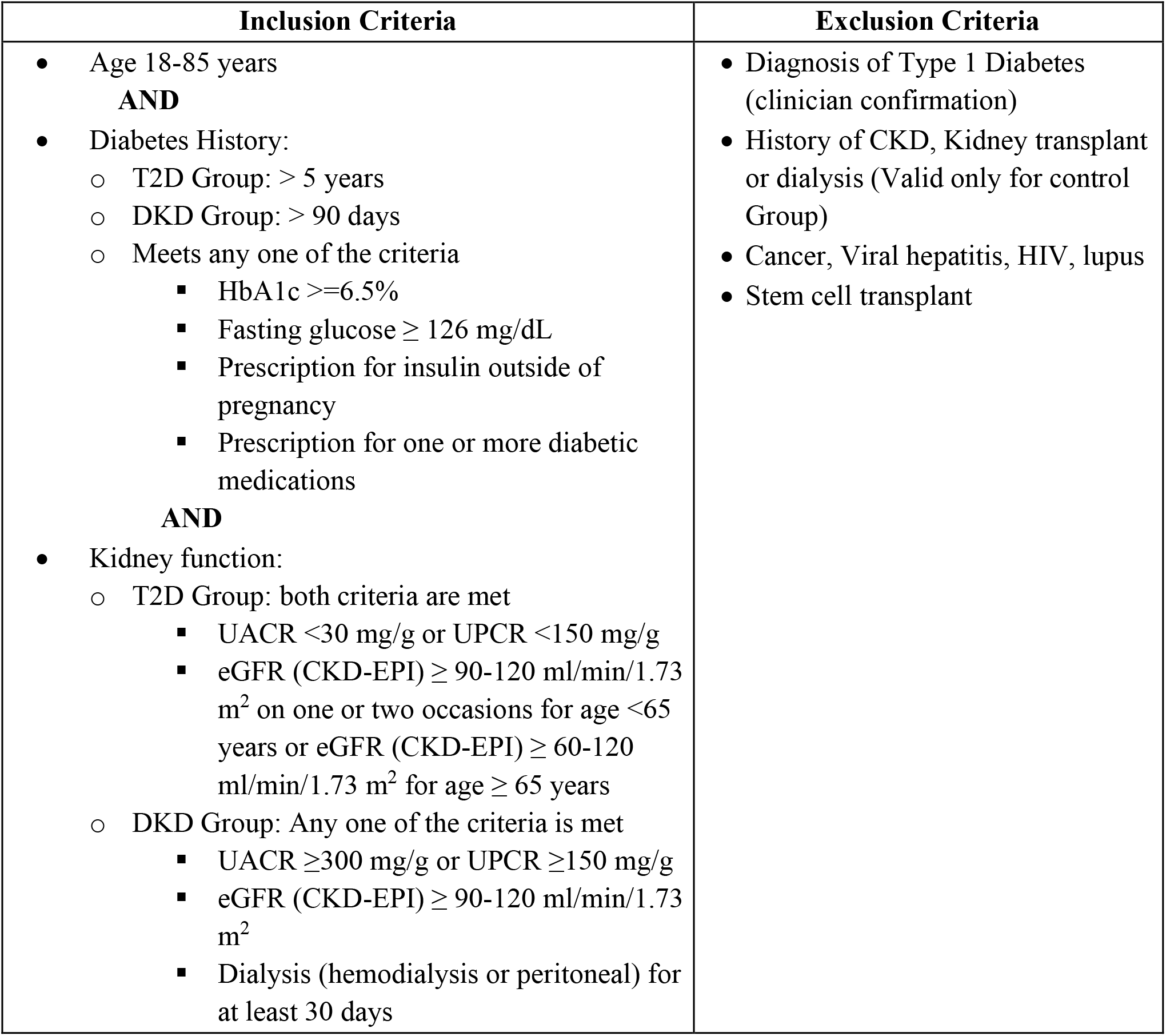
Inclusion and Exclusion criteria for participants recruitment to the study.

**Table 2:** List of study centers enrolled in current study.

| **Site ID** | **Site** | **City** | **# DKD Group** | **# Control Group** | **Total** |
| --- | --- | --- | --- | --- | --- |
| 1 | Arthur Asirvatham Hospital | Madurai | 22 | 21 | 43 |
| 2 | Dr. Gopinath Diabetes Specialty Centre | Madurai | 7 | - | 7 |
| 3 | Hindu Mission Hospital | Chennai | 30 | 9 | 39 |
| 4 | Kidney Center Hospital | Puducherry | 22 | 2 | 24 |
| 5 | Madras Medical Mission Hosp. | Chennai | 15 | 26 | 41 |
| 6 | Madurai Institute of Diabetes and Endocrine Practice and Research | Madurai | 2 | 1 | 3 |
| 7 | Meenakshi Mission Hospital | Chennai | 67 | - | 67 |
| 8 | R.M Diabetes Education & Research | Chennai | 50 | 122 | 172 |
| 9 | SRM Medical College Hospital | Chennai | 23 | 3 | 26 |
|  | Total |  | 238 | 184 | 422 |

In brief, DKD participants were defined as having T2D history of greater than 3 months (at the time of recruitment), eGFR (CKD-EPI) < 60 mL/min/ 1.73m^2^ and UACR > 300 mg/g. T2D participants were defined as having T2D history for more than 5 years (at the time of recruitment) and their kidney was functioning normal as defined by the biochemical test (eGFR (CKD-EPI) =90-120 ml/min/1.73m^2^ for age <65 years or > 60-120 ml/min/1.73m^2^ for age ≥65 years and UACR < 30 mg/g). None of the eligible participants had a history of Type 1 Diabetes Mellitus, any preexisting renal disease, undergone stem cell transplantation, cancer, viral hepatitis, lupus, or nondiabetic kidney disease (NDKD) (Table 1).

All participants were enrolled in the study after they had been briefed up by concerned physician or qualified clinical research coordinator (CRC) and signed the IEC approved informed consent form (ICF) on the day of their regular health visit i.e., screening visit. The goals, potential risks, and advantages of the study were all carefully communicated to the participants in their local language if needed.

Once the interested participants have signed the ICF, blood and urine samples were collected for carrying out the biochemical tests; HbA1c, Serum Creatinine, eGFR (CKD-EPI), Urine Albumin Creatinine Ratio (UACR)/ Urine Protein Creatinine Ratio (UPCR). Additionally, blood samples were collected, and the DNA (Deoxyribonucleic Acid) was extracted and stored for future studies.

Face-to-face interviews were used to collect clinical data in the form of questionnaires covering the demographics, social history, medical history, and family history of the participants. Old laboratory details, case history and concomitant medications were also collected from the participants and digitized. The data digitization was carried out by trained clinical research coordinator using bespoke database platform called “see DISC” developed by Zifo RnD Solutions, Chennai, India. The integrity of the entered data was validated through quality control as well as by the site monitoring visits.

Concomitant Medications were collected, and medical coding was performed using WHODrug 2020 version. ATC Terms & Codes for 4 levels, trade name and drug code are captured as part of medical coding. All the coded terms were reviewed to ensure its medical correctness by another coding specialist or reviewer.

Henceforth, participants belonging to control group will be referred as T2D group/T2D participants while those belonging to case group will be referred as DKD group/DKD participants from this section.

#### Definitions

##### Type 2 Diabetes Mellitus

T2D was diagnosed based on drug treatment for diabetes (insulin or oral hypoglycaemic agents) and/or criteria laid by the WHO consultation report i.e., fasting plasma glucose (FPG) ≥ 126 mg/dl. HbA1c provides a reliable measure of chronic glycemia and correlates well with the risk of long-term diabetes complications, so that it is currently considered as the test of choice for monitoring and chronic management of diabetes. The ADA has recently recommended HbA1c with a cut point 6.5% for diagnosing diabetes as an alternative to fasting plasma glucose (FPG 7.0 mmol/L) (Sherwani et al. 2016)

##### Diabetic Kidney Disease

Participants were considered to have DKD if UACR is increased or if eGFR is reduced in the absence of signs or symptoms of other primary causes of kidney damage. eGFR was considered abnormal if it is less than 60 mL/min/1.73 m^2^ using 2009 CKD-Epidemiology Collaboration (CKD-EPI) creatinine equation (Levey et al. 2009). UACR was considered abnormal if ≥30 mg albumin/g creatinine (mg/g) (Unnikrishnan I et al. 2007).

##### Hypertension

Participants with self-reported hypertension and those who had a systolic blood pressure (SBP) of 140 mmHg and/or diastolic blood pressure (DBP) of 90 mmHg were considered to have hypertension as per National Health Mission Standard Treatment Guidelines (https://nhm.gov.in/images/pdf/guidelines/nrhm-guidelines/stg/Hypertension_full.pdf)

##### Overweight/ Obesity

Obesity was defined using the revised criteria for Asian Indians: underweight: BMI < 18.5 Kg/m2, normal range: BMI 18.5 -22.9 Kg/m2, overweight: at risk: BMI 23.0 - 24.9 Kg/m2, obese I: BMI 25.0 - 29.9 Kg/m2, obese II: BMI ≥ 30 Kg/m2 for both males and females (Kim 2016).

##### Statistical analysis

The study was analyzed by using descriptive statistical analysis using SAS version 9.9. Collected data was analyzed using descriptive statistics including mean, standard deviation (SD) and inferential statistics(p-value) with 5% significance was tested.

## Results

### Recruitment rate and sites

Our initial plan was to recruit 500 participants from 9 sites. However, due to Covid-19 pandemic, the study was cut short to 7 months, and we were able to enroll 482 participants.

Out of 482 participants, 422 (87.6%) met the inclusion criteria while the remaining 60 participants did not meet the criteria. 82.0% of the screen failed participants initially belonged to T2D group, however based on their biochemical results especially eGFR values, they were showing signs of early-stage nephropathy. Majority of the participants were neither smoking nor were taking any abuse substances.

Study participants were recruited from 9 centers that includes hospitals, clinics, and research facilities around Tamil Nadu and Puducherry. 81.8% of the eligible participants were recruited from different sites within Chennai, the fourth largest city in India and capital of Tamil Nadu (Table 2). Among the eligible participants, 71.0% of them were outpatients who had come for routine health checkups and the remaining of them were either visiting for regular dialysis (22.0%) or were hospitalized for various other reasons (7.0%) [Fig.1].

**Fig 1:**
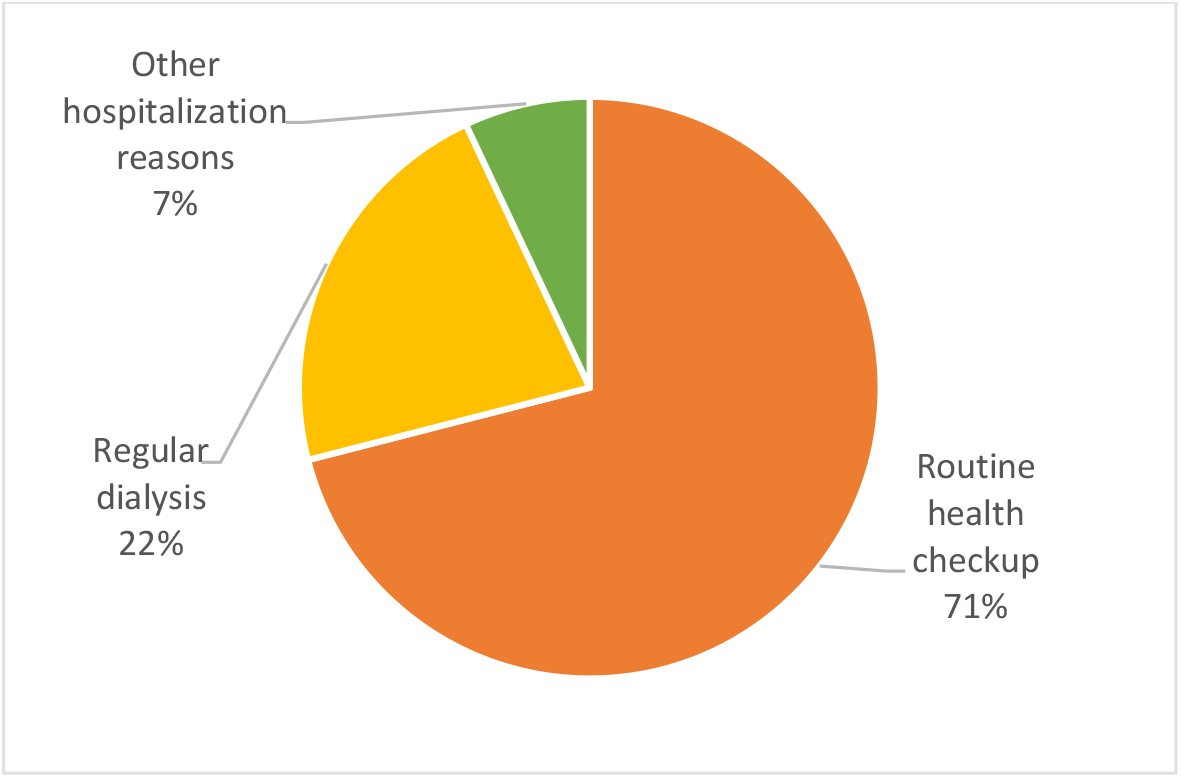
Participants visit status: This figure illustrates the proportion of participants based on the type of visit.

**Fig 2:**
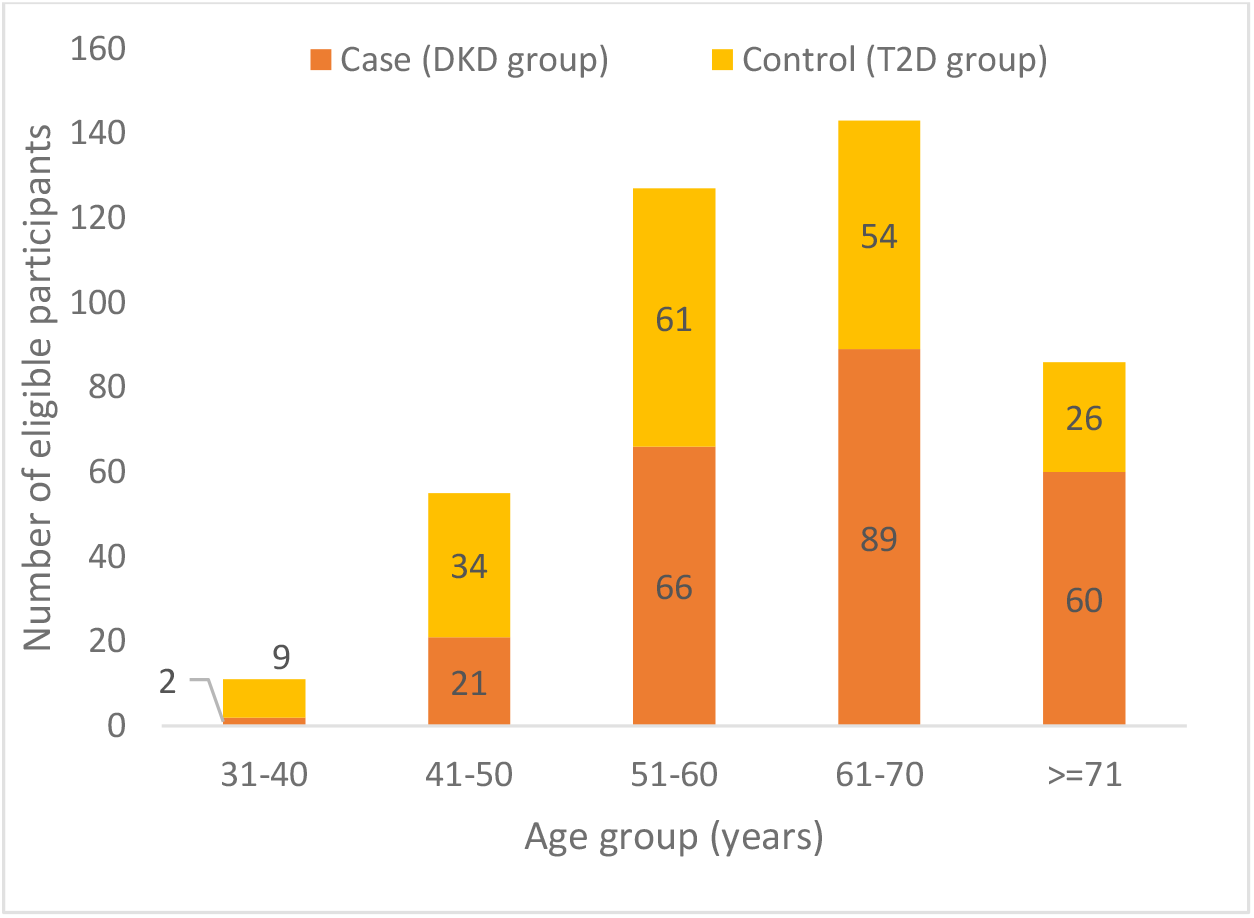
Prevalence of T2DM (Type 2 Diabetes Mellitus) and Diabetic Kidney Disease in various age groups.

Eligible participants were categorized either into DKD group or T2D group based on physician confirmation and biochemical test result. 238 (56.4%) participants belonged to the DKD group (participants having T2D with kidney disease) and the remaining 184 (43.6%) participants formed the T2D group (participants having T2D with healthy kidney).

### Baseline characteristics of the study participants

The mean age of the participants was 61.4±10.5 years and gender wise male were overrepresented in both the groups (65.5% in DKD group and 55.4% in T2D group). Age wise, participants belonging to the DKD group (63.5±9.7 years) were significantly (p value<.0001) older compared to the T2D group (58.6±10.8 years).

The combined mean body mass index (BMI) was 26.3±5.1 kg/m^2^ with 19.4% of them having BMI ≥30 kg/m^2^.Major proportion of them belonged to the DKD group (57.3%). Obesity was more prevalent in women (59.8%) when compared to men (40.2%). Proportion of obese male was higher in DKD group (69.7%) than T2D group while it was evenly split in both the groups for female participants.

Hypertension was reported in 229 (54.3%) individuals based on the blood pressure measurements taken on the day of screening, with major proportion belonging to the DKD group. The mean systolic blood pressure was 139.0±21.8 mmHg and the mean diastolic blood pressure was 80.0±10.8 mmHg. The mean systolic blood pressure in DKD group was 144.0±21.8 mmHg and diastolic blood pressure was 81±11.3 mmHg, while mean systolic blood pressure in T2D group was 132±19.7 mmHg and diastolic blood pressure was 78.0±9.9 mmHg, respectively. Diastolic blood pressure was significantly different between the two groups but not as significant as systolic blood pressure [Table 3].

**Table 3:** Baseline characteristics of participants with T2D with normal kidney function (control group) and T2D with kidney disease (DKD group).

| Parameters | Overall<br>(N=422) | DKD<br>Group<br>(N=238) | T2D<br>Group<br>(N=184) | p-Value |
| --- | --- | --- | --- | --- |
| **Age, years, mean±SD** | 61.4±10.5 | 63.5±9.7 | 58.6±10.8 | <.0001*** |
| **Gender, n (%)** |  |  |  |  |
| Female | 164(38.9) | 82(34.5) | 82(44.6) | 0.0346* |
| Male | 258(61.1) | 156(65.5) | 102(55.4) | 0.0346* |
| **Weight, kg, mean±SD** | 68.2±13.9 | 67.5±15.0 | 69.2±12.2 | 0.1991 |
| **BMI, kg/m², mean±SD** | 26.3±5.1 | 26.2±5.3 | 26.5±4.8 | 0.6327 |
| **BMI kg/m², n (%)** |  |  |  |  |
| Underweight <18.5 | 5(1.2) | 5(2.1) | 0 | 0.0479* |
| Normal 18.5 – 24.9 | 177(41.9) | 98(41.2) | 79(42.9) | 0.7166 |
| Overweight 25-29.9 | 158(37.4) | 88(37.0) | 70(38.0) | 0.8220 |
| Obese ≥30 | 82(19.4) | 47(19.7) | 35(19.0) | 0.8517 |
| **Systolic Blood Pressure, mm Hg, mean±SD** | 139.9±21.8 | 144.0±21.8 | 132.0±19.7 | <.0001*** |
| **Diastolic Blood Pressure, mm Hg, mean±SD** | 80.0±10.8 | 81.0±11.3 | 78.0±9.9 | 0.0204* |
| **Blood Pressure mm Hg, n (%)** |  |  |  |  |
| Normal (SBP<120 and DBP<80) | 52(12.4) | 14(5.9) | 38(20.7) | <.0001*** |
| Elevated (SBP 120-139 & DBP 80-89) | 141(33.5) | 64(26.9) | 77(41.8) | 0.0012** |
| Hypertension stage 1 (SBP 140-159 & DBP 90-99) | 137(32.5) | 88(37.0) | 49(26.6) | 0.0244* |
| Hypertension stage 2 (SBP ≥160 or DBP ≥100) | 92(21.9) | 72(30.3) | 20(10.9) | <.0001*** |
| **Pulse Rate, beats/min, mean±SD** | 82±10.5 | 83±10.2 | 80±10.6 | 0.0022** |
| **Age of onset of T2D, years, mean±SD** | 46±10.4 | 47±10.5 | 44±10.1 | 0.0011** |
| **Type 2 Diabetes duration, years, mean±SD** | 15±7.5 | 16±8.1 | 14±6.5 | 0.0200* |
| **Type 2 Diabetes duration, n (%)** |  |  |  |  |
| <=5 Years | 28(6.7) | 17(7.1) | 11(6.0) | 0.6336 |
| 5-10 Years | 102(24.2) | 60(25.2) | 42(22.8) | 0.5705 |
| 10-20 Years | 203(48.2) | 100(42.0) | 103(56.0) | 0.0044** |
| 20-30 Years | 75(17.8) | 51(21.4) | 24(13.0) | 0.0255* |
| 30-40 Years | 12(2.9) | 8(3.4) | 4(2.2) | 0.4668 |
| 40-50 Years | 1(0.3) | 1(0.4) | 0 | 0.3787 |
| >50 Years | 1(0.3) | 1(0.4) | 0 | 0.3787 |
| **Medical History, n (%)** |  |  |  |  |
| Cardiovascular disease | 23(5.5) | 10(4.2) | 13(7.1) | 0.1988 |
| Coronary artery disease | 59(14.0) | 42(17.6) | 17(9.2) | 0.0135* |
| Diabetic retinopathy | 22(5.2) | 20(8.4) | 2(1.1) | 0.0008** |
| Heart failure | 3(0.7) | 2(0.8) | 1(0.5) | 0.7189 |
| Hypertension | 272(64.5) | 192(80.7) | 80(43.5) | <.0001*** |
| Peripheral vascular disease | 10(2.4) | 8(3.4) | 2(1.1) | 0.1277 |
| **HbA1c, %, mean±SD** | 7.7±1.7 | 7.5±1.7 | 8.0±1.7 | 0.0019** |
| **HbA1c, n (%)** |  |  |  |  |
| Normal (<5.7%) | 17(4.1) | 17(7.1) | 0 | 0.0002*** |
| Prediabetic (5.7% to 6.5%) | 62(14.7) | 43(18.1) | 19(10.3) | 0.0259* |
| Diabetes (>=6.5) | 343(81.3) | 178(74.8) | 165(89.7) | 0.0001*** |
| **Serum Creatinine, mg/dL, mean±SD** | 2.6±2.8 | 4.1±3.0 | 0.8±0.1 | <.0001*** |
| **UACR, mg/g, mean±SD** | 842.6±263<br>6.8 | 1538.6±343<br>8.5 | 16.9±7.3 | 0.0004*** |
| **UACR, mg/g, n (%)** |  |  |  |  |
| Normal to Mildly increased (<30) | 70(16.6) | 11(4.6) | 59(32.1) | <.0001*** |
| Moderately increased (30-299) | 19(4.6) | 19(8.0) | 0 | <.0001 |
| Severely increased (≥300) | 40(9.5) | 40(16.8) | 0 | <.0001 |
| **eGFR (CKD-EPI), mL/min/1.73m<sup>2</sup>, mean±SD** | 56.1±38.3 | 26.7±22.1 | 94.1±13.4 | <.0001*** |
| **eGFR (CKD-EPI), mL/min/1.73m<sup>2</sup>, n (%)** |  |  |  |  |
| G1 Normal or High ≥90 | 146(34.6) | 5(2.1) | 141(76.6) | <.0001*** |
| G2 Mildly decreased (60-89) | 50(11.9) | 7(2.9) | 43(23.4) | <.0001*** |
| G3a Mildly to Moderately Decreased (45-59) | 46(11) | 46(19.3) | 0 | <.0001 |
| G3b Moderately to Severely Decreased (30-44) | 36(8.6) | 36(15.1) | 0 | <.0001 |
| G4 Severely Decreased (15-29) | 37(8.8) | 37(15.5) | 0 | <.0001 |
| G5 Kidney Failure (<15) | 107(25.4) | 107(45.0) | 0 | <.0001 |

Note: \*\*\* $p \leq 0.0005$ , \*\* $p \leq 0.005$ , \* $p \leq 0.05$ . When a value in either the DKD or T2D group is zero, the p value is not considered to be significant.

The mean age of onset of T2D was seen to be 46.0±10.4 years with the mean duration of 15.0±7.5 years [Table 3]. Notably, 48.1% (203) participants were diagnosed with T2D for a duration of 10-20 years [Table 3]. 46.0% of the total eligible participants suffering from T2D were between the age of 61-70 years.

A mean HbA1c of 7.7±1.7% was seen in eligible participants. The mean HbA1c among T2D participants (8.0±1.7%) was statistically higher (p=.0019) compared to DKD participants (7.5±1.7%).

The mean serum creatinine concentration at the time of screening was 2.6±2.8 mg/dL, and the UACR concentration was 842.6±2636.8 mg/g [Table 3], respectively.

### DKD (Diabetic Kidney Disease) and its related laboratory parameters

Diabetic Kidney Disease (DKD) was more prevalent among the DKD group having age above 50 years. Age wise distribution of DKD participants having DKD was 0.8%, 8.8%, 13%, 27.7%, 37.4% and 25.2%, respectively for the age groups 21-30, 31-40, 41-50, 51-60, 61-70 and >70 years [Fig 1].

Renal functions were assessed at the time of screening using either Urine Albumin Creatinine Ratio (UACR) or the Urine Protein Creatinine Ratio (UPCR) and / or their Estimated Glomerular Filtration Rate (eGFR) CKD-EPI values. UACR and UPCR data was collected from 129 and 94 participants in both groups, however, in case of DKD participants who were on routine dialysis, UACR / UPCR values was not collected.

Among 129 participants whose UACR value was collected, 70 (54.3%) had normal to mildly increased albuminuria (A1; normoalbuminuric; UACR <30 mg/g), 19 participants (14.7%) had moderately increased albuminuria (A2; microalbuminuria; UACR 30–299 mg/g) and 40 participants (31.0%) had severely increased albuminuria (A3; macroalbuminuria; UACR >300 mg/g) [Table 3]. 79 of 94 participants with UPCR value felled in normal range (<150 mg/g) and belonged to T2D group while rest of the participants belonged to DKD group.

eGFR was measured for all the participants. 146 participants fell under G1 category (>=90 mL/min/1.73m^2^), 50 participants had eGFR values fall under G2 category (60-89 mL/min/1.73m^2^), 46 participants had values in G3a category (45-59 mL/min/1.73m^2^), 36 participants in G3b category (30-44 mL/min/1.73m^2^), 37 participants had eGFR value falling in G4 category (15-29 mL/min/1.73 m^2^) and for remaining 107 participants eGFR value were in G5 category (<15 mL/min/1.73m^2^) and they had kidney failure, respectively [Table 3]. Major proportion of the DKD participants (107) fell into G5 category (<15 mL/min/1.73 m^2^), and 85 (79.4%) of them were undergoing dialysis with multiple (>1) sessions per week. All the T2D participants belonged to either G1 or G2 category [Table 4].

**Table 4:**
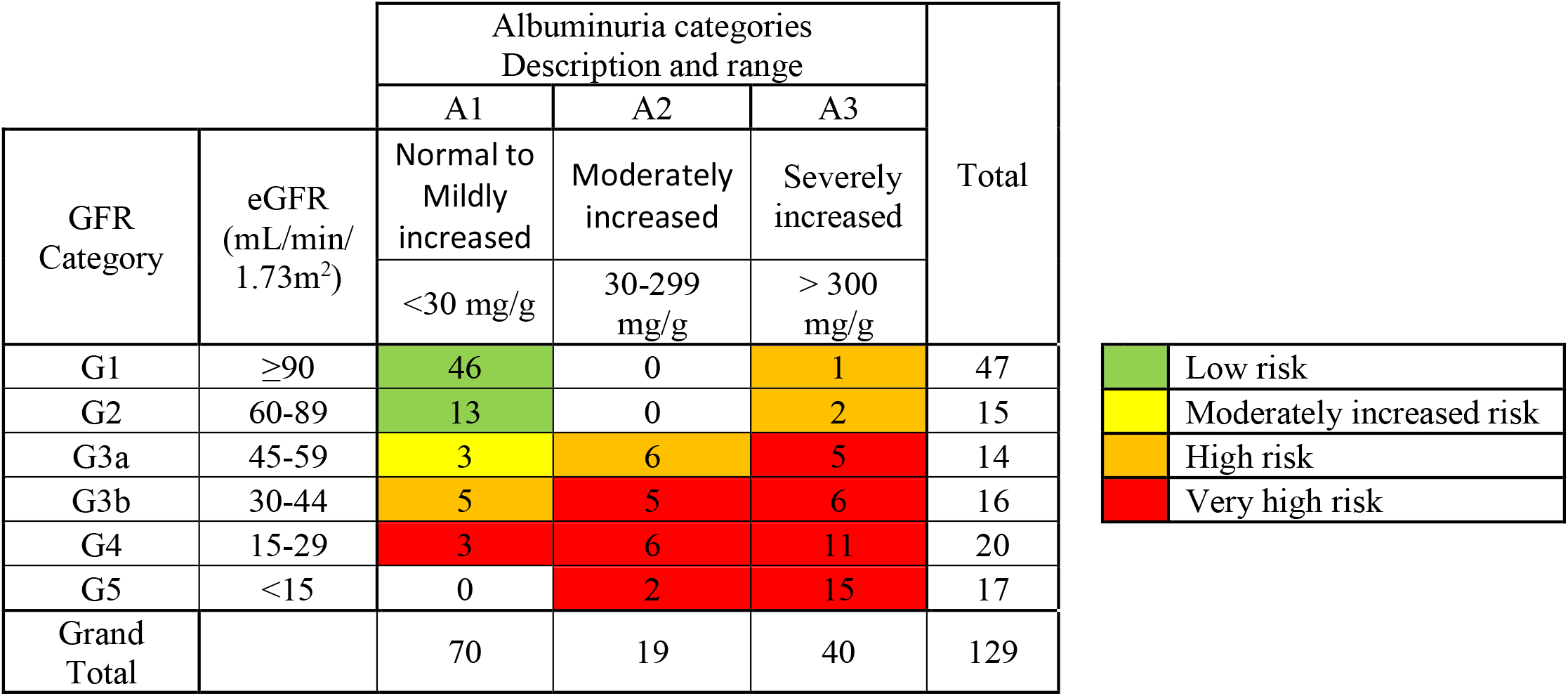
Prevalence of CKD (chronic kidney disease) in eligible participants based on UACR and eGFR according to KDIGO classification system.

According to the Kidney Disease Improving Global Outcomes (KDIGO) guidelines 2021, we categorized participant’s kidney function based on eGFR and UACR values (Ronco et al.) Only for 129 participants both eGFR and UACR values were present. 51.93 % of the participants belonged to either high risk or very high-risk category of kidney disease [Table 4].

### Prevalence of comorbid conditions and complications

Participants had other comorbidities also along with T2D that includes hypertension, diabetic retinopathy, coronary artery disease, cardiovascular disease, peripheral vascular disease, and heart failure. [Table 3].

The number of comorbidities that participants had in addition to T2D ranged from 1 to 5. Other than T2D, 210 eligible participants had only one additional comorbidity, 70 had two, and 10 had three comorbidities, respectively. Two of the DKD participants had more than three comorbidities.

64.5% of the participants had history of hypertension along with T2D and majority of them belonged to DKD group. 160 DKD participants and 69 T2D participants fell into hypertension stage 1 (SBP 140-159 mmHg or DBP 90-99 mmHg) and hypertension stage 2 (SBP >160 or DBP >100 mmHg) categories, respectively. [Table 3].

Prevalence of retinopathy was reported in 22 of the participants, with 90.9% of them falling in the DKD group. Likewise, coronary artery disease also was found more often in DKD group (42 of 59 participants) [Table 3].

### Family History of Diabetes

81 participants in our study had reported either one of their first-degree relatives with family history of either T2D or DKD. A total of 55 participants had diabetic family history with at least one of the parents suffering from T2D and it was overrepresented (54.5%) in the T2D group. There were 11 participants whose both the parents had history of T2D, and they were equally represented in both the groups (T2D group n =6, DKD group n = 5). There were 12 participants whose one or both parents were affected with diabetic kidney disease and was equally distributed among both groups (T2D group n =4, DKD group n = 4).

### Diabetic medications

For 400 participants, information on antihyperglycemic treatments was available. 306 (76.5%) participants were on insulin prescription. Apart from insulin, the participants were also taking different class of antihyperglycemic medications. The majority of the participants were treated with Biguanides, however significantly lower proportion of patients in the DKD group received metformin compared to those without DKD (45% vs 55%). Dipeptidyl peptidase 4 (DPP-4) inhibitors were the next commonly used medication (38% in DKD group vs 62% in the T2D group) followed by sulphonylureas (44% in DKD group vs 56% in the T2D group) [Table 5]. Insulin was prescribed regularly among participants having chronic kidney disease and cardiovascular disease, while sodium-glucose cotransporter 2 inhibitors and Glucagon Like Peptide 1 receptor agonists were rarely prescribed.

**Table 5:** Antihyperglycemic agents used by the participants.

| Therapy | DKD(n) | Control(n) | Total(n) |
| --- | --- | --- | --- |
| Insulin | 209 | 97 | 356 |
| Biguanides | 86 | 165 | 252 |
| Dipeptidyl peptidase 4 (DPP-4) inhibitors | 89 | 146 | 235 |
| Sulfonylureas | 102 | 128 | 230 |
| Sodium-glucose co-transporter 2 (SGLT2) inhibitors | 28 | 82 | 110 |
| Combinations of oral blood glucose lowering drugs | 33 | 43 | 76 |
| Alpha glucosidase inhibitors | 25 | 30 | 55 |
| Other blood glucose lowering drugs, excl. insulins | 17 | 33 | 50 |
| Glucagon-like peptide-1 (GLP-1) analogues | 11 | 5 | 16 |

## Discussion

In this retrospective study, we provide an insight into the clinical and epidemiological characteristics of T2D and DKD individuals from southern part of India, primarily Tamil Nadu. To understand the associated risk factors in the development of kidney disease, we compared the baseline and other clinical parameters of the participants belonging to DKD group (T2D participants with kidney disease) and T2D group (T2D participants with healthy kidney).

We found male sex, increasing age, longer duration of T2D, and hypertension as the major risk factors of DKD. In addition, we found a higher risk of developing additional comorbidities such as diabetic retinopathy, coronary artery disease among the DKD individuals.

In our study, we found higher prevalence of T2D and DKD among men than women. The ratio being 2:1 for DKD participants and 11:4 for T2D participants, respectively and was statistically significant. Our findings align with earlier studies that have reported higher prevalence of T2D and DKD among Indian men compared to women (Kumar et al. 2022) (Ramachandran et al. 1988) (Yu et al. 2012). Additionally, studies like UKPDS 74 study, where Retnakaran and his coworkers found male sex as a risk factor for DKD. They also reported that males were more likely to follow an albuminuric pathway leading to decline in eGFR value whereas females were more likely to follow non-albuminuric pathway (Retnakaran et al. 2006). In contrast, very few studies have reported female sex as a risk factor of DKD (Roy et al. 2021).

Many studies have shown increasing age as a key risk factor for the development of DKD (Metsärinne et al., 2015), (Tewari et al., 2021). As people age (>60 years), Roy et al. showed that renal function (eGFR value) decreased in both diabetes and non-diabetic individuals (Roy et al., 2021) and is in line with the findings of our study in both the groups. We observed that in T2D individuals eGFR value declines with age, while in DKD individuals eGFR value was fluctuating with increasing age.

Studies have reported that longer duration of diabetes increases the prevalence rate of albuminuria. In a study it has been reported, if duration of diabetes was less than 4 years the prevalence rate of albuminuria is around 20.9% which increases to 54.1% for duration greater than or equal to 20 years (Reutens 2013). Another study involving a large cohort of 27,029 persons with T2D showed that elderly people and those who had diabetes for a longer duration were having 49.0% risk of developing DKD (Goderis et al. 2013) (de Cosmo et al. 2016). On lines - to the earlier study findings, we found that longer duration of diabetes plays a role in the development of DKD. Many of our study participants having diabetic history of more than 10 years are suffering from DKD which is in line with earlier study reports. T2D participants having diabetic duration of greater than 10 years showed early stages of nephropathy and were falling under G2 category of eGFR classification i.e., having kidney damage with a mild decrease in their glomerulus filtration rate (GFR).

The World Health Organization (WHO) defines overweight and obesity as abnormal or excessive fat accumulation that presents a risk to health (WHO, 2016a). In our study, participants having BMI greater than 30 kg/m^2^ were considered obese (Kim 2016)(WHO, 2016a). Agrawal et al. in his 2017 study reported obesity as one of the risk factors associated with kidney disease (Agrawal et al. 2017) however in current study no relation could be established between obesity and DKD. Based on our results, obesity cannot be considered as a significant factor for predicting kidney disease. In 2013, Maric-Bilkan and his coworkers studied the mechanism by which obesity may lead to end-stage renal disease (ESRD) and reported obesity as one of the driving forces leading to increase in prevalence of T2D and hypertension which indirectly paves the way for DKD (Maric-Bilkan 2013).

The Framingham study cohort showed that BMI could predict reduced kidney function regardless of the presence of comorbidities like diabetes or hypertension (Foster et al. 2008), (Maric-Bilkan 2013). However, in respect to BMI, the current study is in lines with Thakkar et al. study, where the author failed to show any correlation between BMI and DKD among newly diagnosed participants with Diabetes Mellitus (Thakkar et al. 2011). Janssen et al., evaluated the predictive power of BMI in assessing the obesity related comorbidities in his study and reported that BMI may not be a robust parameter for predicting kidney disease as BMI cannot differentiate between body muscle and body fat. Since BMI does not account for the difference between the body muscle and body fat, it might not be used as a dependable indicator of reduced eGFR or decline in renal function, particularly among T2D people (Janssen et al. 2004).

We saw poor glycaemic control among the DKD participants compared to T2D group and this correlation was statistically significant (p value=<.0001). In line with our study, Mir et al. in 2019 also reported statistically significant positive correlation between prevalence of DKD and increase of HbA1c (Mir et al. 2019). Additionally, few studies have reported HbA1c value greater than 8% being associated with higher incidence of kidney disease by increasing the albumin concentration (Debbarma et al. 2015) (Limkunakul et al. 2019). Study by Viberti in the newly diagnosed T2D patients reported a decrease in albumin excretion with improved glucose levels (Viberti et al. 1979). A great deal of evidence reports that good control of blood glucose levels prevents DKD development (Muskiet et al. 2014).

Family history has been reported as one of the independent risk factors for the development of T2D in many studies (Anjana et al. 2017) (Scott et al. 2013). In our study, 19% of the total participants had reported family history of T2D among their first-degree relatives. Among those participants, we observed the incidence of association of family history of T2D was lower in participants with DKD compared to participants with T2D. In 2021, Roy et al., had reported similar findings about the association of family history of diabetes significantly among the group of participants with T2D and without DKD (Roy et al. 2021). In 2017, Geetha A et al., studied the impact of family history of diabetes among the T2D patients in Kancheepuram district at Tamil Nadu. They showed that participants with positive family history of diabetes were more prone to early onset of diabetes and development of other complications (A. et al. 2017). Many T2D studies have shown that family history plays a major role in its progression of T2D to the next generation (A. et al. 2017), (Anjana et al. 2017). Scott in his study has shown that the greatest risk of developing T2D was among those participants who had biparental history of T2D (Scott et al. 2013). Wang in his study involving young participants suffering from diabetes has reported rapid decline in eGFR was significantly higher in participants having diabetic family history especially first-degree relatives. (Wang et al. 2019). In the current study, we do not see a decline in eGFR value based on family history probably due to small sample size. Moreover, many studies have linked the family history and risk of T2D to both genetic and shared environmental components among the family members, but the precise factors accounting for this increase in risk are poorly understood (Scott et al. 2013). We would like to extend the scope of this study to understand the influence of genetics in T2D by genotyping of the samples collected from the participants.

Several studies have reported higher frequencies of association of hypertension, hyperlipidemia, coronary artery disease, cerebrovascular accidents among the DKD participants. (Roy et al. 2021). In line with these studies, we observed an association of several comorbid medical conditions such as hypertension, coronary artery disease and diabetic retinopathy significantly higher in our DKD participants compared to T2D participants. Many studies have reported hypertension as an important independent risk factor for DKD (Tapp et al. 2004) (Agrawal et al. 2017) (Colosia et al. 2013) (Tedla et al. 2011). In line with the previous study reports, hypertension was significantly higher in our participants with DKD when compared to T2D participants (p <.0001). Additionally, studies have shown the existence of hypertension among the T2D participants prior to kidney disease (van Buren and Toto 2011). Study by Do Carmo have reported that hypertension combined with diabetes and obesity contributes to progression of renal injury and DKD (do Carmo et al. 2009). Kidney Disease Outcomes Quality Initiative (KDOQI) guidelines emphasize the necessity of strict blood pressure management in treating DKD, with a suggested blood pressure target of less than 130/80 mmHg (van Buren and Toto 2011). Further, several studies have shown clear renal protection with respect to slowing progression of nephropathy in patients with T2D via blood pressure lowering (Grossman and Messerli 2011) (Heerspink and de Zeeuw 2011).

Early diagnosis of kidney disease is a necessity of time, and it will help to control the progression of the disease, extend the life expectancy thereby reducing the humanistic and economic load (Hussain et al. 2021b). For the diagnosis of DKD, the most used available markers in clinical practice are albuminuria (UACR) and serum creatinine (eGFR) (Hussain et al. 2021b). UACR is highly recommended by ADA guidelines for early diagnosis of DKD (Stempniewicz et al. 2021) however Retnakaran et al., reported that UACR fails to identify advancing renal disease to its final stages (Retnakaran et al., 2006). Branten et al showed that serum creatinine predicts DKD only if the damage is severe (Branten et al. 2005). Such studies outline the importance of monitoring of both eGFR and UACR twice annually in patients with diabetes(Stempniewicz et al. 2021). In our study, we observed 2 DKD participants who had normal eGFR value but when considering UACR, the value was abnormal (>300 mg/g), conversely, we also found 11 DKD participants had normal UACR value but eGFR value was less than normal range, underlying the importance of regular measurement of both markers in especially among DKD individuals on regular intervals yearly.

We observed that more than half of the study participants in both groups were managed by one or more diabetic medications. In our study, a considerable number of DKD participants were prescribed with insulin rather than oral diabetic medications and these results were in-line with the study conducted by Roy et al., in 2021 (Roy et al. 2021). Among the various classes of medications, the majority of our T2D participants were treated with metformin (Biguanides) compared to DKD participants. Shaw et al., had proposed pragmatic eGFR limits to guide metformin prescribing in patients with renal impairment. It has been reported that CKD stage 4 or greater should be an absolute contraindication to metformin, while CKD stage 3 should alert clinicians to consider other risk factors before initiating or continuing treatment with metformin (Shaw et al. 2007). However, metformin has been used safely, without causing hypoglycemia in patients with prediabetic hyperglycemia (Knowler WC et al.,2002). Next commonly used drugs were DPP-4 inhibitors and Sulphonylureas. These results were in line with other study reports which shows metformin as a common drug used among the diabetic patients in India followed by other drug categories (Roy et al. 2021).

Being a retrospective study, our study has few limitations. Since we did not have direct interaction with the participants and all the medical and sociodemographic information were shared by the site-specific clinical research coordinators, hence it is possible that certain information, such as disease duration, medical and family history, could not been reliably ascertained. Another limitation was that the COVID pandemic restricted us from recruiting the number of participants planned in our study and we were able to enroll less than 500 participants, which does not represent the true subset population of a region. An observational study with small sample size and possible selection bias, findings of our study need to be carefully reviewed before considering it generalizable to a general population with T2D and DKD. The key strength of our study was the availability of participants who were recruited from specialists treating T2D and DKD and they were categorized into two groups based on stringent eligibility criteria. Additionally, all the data entered by the site team was reviewed and had undergone source data verification at site by the study monitoring team.

## Data Availability

All data produced in the present study are available upon reasonable request to the authors

## Acknowledgement

We would like to thank clinical research coordinators of the enrolled clinical sites whose patience, conscientiousness and creativity made this study successful and helping in digitizing the participants medical data.

We would like to express our sincere gratitude to Zifo RnD Solutions, Chennai, India for providing financial support to conduct the study.

We acknowledge our institute’s data management and statistical team for their immense support in providing and analysing the data. Statistical work was carried out by Sangli Therasanathan, Aparna Bhide. Data management and validation was carried out by Surendhar Venkatesan, Dhamodharan R, Georgeena Sheela Jayasheelan. We would like to thank all the persons involved in this study and helped us in different ways.

## Notes

### Competing Interest Statement

The authors have declared no competing interest.

### Funding Statement

This study did not receive any funding

### Author Declarations

Ethics committee of Hindu Mission Hospital, Chennai gave ethical approval for this work. Ethics committee of Madras Medical Mission Hospital, Chennai gave ethical approval for this work. Ethics committee of SRM Medical College Hospital & Research Centre, Chennai gave ethical approval for this work. Ethics committee of Arthur Asirvatham Hospital gave ethical approval for this work. Ethics committee of Meenakshi Mission Hospital & Research Centre gave ethical approval for this work. Ethics Committee of Jehangir Clinical Developmental Centre (JCDC) Ethics Committee pvt. ltd. gave ethical approval for this work.

### Summary of Updates

Updated author's name to full name. Made few cosmetic changes in the references.

